# A multicenter, randomized, blinded, controlled clinical trial investigating the effect of a novel infant formula on the body composition of infants: INNOVA 2020 study protocol

**DOI:** 10.1101/2022.08.31.22279449

**Authors:** Francisco Javier Ruiz-Ojeda, Julio Plaza-Díaz, Javier Morales, Ana Isabel Cristina de la Torre, Antonio García-García, Carlos Nuñez de Prado, Cristóbal Coronel, Cyntia Crespo, Eduardo Ortega, Esther Martín-Pérez, Fernando Ferreira, Gema García-Ron, Ignacio Galicia, María Teresa Santos-García Cuéllar, Marcos Maroto, Paola Ruiz, Raquel Martín, Susana Viver-Gómez, Ángel Gil

**Author notes:** **Corresponding author:** Prof. Angel Gil, Department of Biochemistry and Molecular Biology II, School of Pharmacy, University of Granada, Campus de Cartuja sn, 18071, Granada, Spain, e-mail addresses. Equally contributed (Francisco Javier Ruiz-Ojeda, Julio Plaza-Díaz).

## Abstract

**Background:** Breastmilk is the ideal food for infants and exclusive breastfeeding is recommended. In the clinical trial aimed to evaluate a new starting formula on weight gain of infants up to 6 and 12 months. The novel formula was compared with a standard formula and breastfeeding, the latter being used as the reference method.

**Methods:** 210 infants (70/group) were enrolled in the study, and completed the intervention until 12 months of age. For the intervention period, infants were divided into three groups: group 1 received the formula 1 (Nutribén Innova®1 or INN), with a lower amount of protein, and enriched in α-lactalbumin protein, and with double amount of docosahexaenoic acid (DHA)/ arachidonic acid (ARA) than the standard formula; it also contained a thermally inactivated postbiotic (*Bifidobacterium animalis* subsp. *lactis*, BPL1™ HT). Group 2 received the standard formula or formula 2 (Nutriben® or STD) and the third group was exclusively breastfed for exploratory analysis. During the study, visits were made at 21 days, 2, 4, 6, and 12 months of age, with ± 3 days for the visit at 21 days of age, ± 1 week for the visit at 2 months, and ± 2 weeks for the others.

**Discussion:** The findings of this study will provide evidence regarding the beneficial health effects of having a novel starting infant formula with reduced levels of protein, enriched in α-lactalbumin, and increased levels of DHA and ARA, and containing a postbiotic, compared with infants fed standard formula.

**Trial registration:** The trial was registered with Clinicaltrial.gov (NCT05303077) on March 31, 2022, and lastly updated on April 7, 2022.

## Background

Breastmilk is the ideal food for infants and exclusive breastfeeding is recommended for the first 6 months of life [1] due to its link to lower infant morbidity and mortality in both developing [2] and developed countries [3, 4]. Importantly, breastfeeding not only benefits child health and development but also holds benefits for maternal health [5, 6]. Indeed, human milk provides all nutrients to support infant growth and development as well as immuno-protection during the critical period of his immature immune system [7]. Moreover, breast milk contains functional nutrients that help provide the microenvironment for gut protection and maturation [8].

Despite the well-known benefits of human milk, for some mothers, breastfeeding is difficult or even impossible, which can result in infants experiencing weight loss. In such situations, formula milk can be used to supplement the infant’s nutritional needs [7]. Even for mothers who can breastfeed increases in maternal employment and a lack of paid parental leave may be causing women to opt for alternative feeding practices [9].

All formulae intended for infants must be safe and suitable to meet the nutritional requirements and promote the growth and development of infants born at term when used as a sole source of nutrition during the first months of life, and when used as the principal liquid element in a progressively diversified diet after the introduction of appropriate complementary feeding [10].

Several international organizations have developed guidelines for the composition of infant formulas. The Codex Alimentarius Commission, part of both the Food and Agriculture Organization of the United Nations (FAO) and the World Health Organization (WHO), has developed standards on infant formula, which are permanently revised [11]. In addition, the European Society for Pediatric Gastroenterology, Hepatology and Nutrition (ESPGHAN) arrived at recommendations on the compositional requirements for a global infant formula standard [12]. Besides, in European countries, the Commission Delegated Regulation (EU) 2016/127 of 25 September 2015 supplementing Regulation (EU) No 609/2013 of the European Parliament and of the Council regulates the specific compositional and information requirements for infant formula and follow-on formula [13]. In the United States, the Food and Drug Administration (FDA) provides strict nutritional and safety standards for infant formulas [14].

Rapid growth and enhanced weight gain during the early years of life are associated with higher body mass index (BMI) and obesity in adulthood [15]. Using infant formula with a relatively high protein intake has been shown to exacerbate the risk of childhood obesity [16]. Indeed, infant formula within the lowest adequate range of protein intake should be clinically evaluated to test to what extent reducing the protein content would result in a more adequate growth velocity.

Modifications to infant formulas are continually being made as the components of human milk are characterized and as the nutrient needs of diverse groups of infants are identified [17]. In the last, 20 years, formulas supplemented with long-chain polyunsaturated fatty acids (LC-PUFA) in amounts similar to those in human milk have become available worldwide. However, new EU regulation [13] indicates that infant formulas should contain between 20 and 50 mg of docosahexaenoic acid (DHA) per 100 kcal (0.5-1% of total fatty acids, higher than in human milk and most commercially available infant formulas) without the need to include also arachidonic acid (ARA). This new regulation has aroused a great deal of controversy, as there is no scientific evidence on its relevance and safety for healthy infants for healthy children. Indeed, international expert consensuses recommend the use of an infant formula that provides DHA at levels of 0.3% to 0.5% by weight of total fat content and with a minimum level of ARA equivalent to the DHA content [18, 19]. Hence, the supplementation of infant formulas with both DHA and ARA in appropriate amounts should be clinically tested.

On the other hand, the use of prebiotics, probiotics, synbiotics, postbiotics, parabiotics and paraprobiotics in infant formulas has increased in the last few years. However, evidence for the health effects of such enrichment in early childhood remains inconclusive [20]. The probiotic strain *Bifidobacterium animalis* subsp. *lactis*, BPL1™, has demonstrated its functionality in preclinical and clinical studies in reducing visceral fat levels in obese individuals [21-23] as well as its ability, even in its inactivated form (postbiotic), to modulate the microbiome [24]. However, until now any study has addressed its potential effects on infants.

As current infant formulas incorporate novel ingredients to partly mimic the composition of human milk, the safety and suitability of each specific infant formula should be tested by clinical evaluation in the target population. Indeed, the present study aimed to report the design and methodology of a randomized double placebo-controlled clinical trial consisting of the evaluation of a novel infant formula with reduced content of total protein and qualitatively modification of the proportion of casein to whey protein ratio of about 70/30 by increasing the content of α-lactalbumin, increased levels of both AA and DHA, and containing a postbiotic (thermally inactivated BPL1™ HT, which is a trademark owned by ADM Biopolis S.L., Spain, registered in the European Union, the USA and other countries), in comparison with standard infant formula. An exclusively breastfed population was followed-up as a reference. The safety, tolerance, and effects on growth and incidence of major acute infectious diseases and changes in intestinal microbiota were evaluated for 6 months, and until 12 months after the introduction of complementary feeding.

## Methods

### Research hypothesis

The study aims to test the following null hypotheses:

1. A novel formula (Formula 1 Nutribén Innova®1 or INN) will lead to similar weight gain as children fed the standard formula (Formula 2 Nutriben® or STD).
2. Infants fed the novel formula 1 will exhibit additional health benefits over infants fed the STD formula 2.

### Objectives

#### Primary objective

The main objective of this study was to evaluate the weight gain in children fed with a new starting infant formula INN for 12 months, with a low level of total protein, enriched in α-lactalbumin protein to get a proportion of whey proteins to caseins of 70/30, as well as enriched levels of docosahexaenoic acid (DHA) and arachidonic acid (ARA) (about 2 times than STD infant formula), and supplemented with the postbiotic *Bifidobacterium animalis* subsp. *lactis* (BPL1™ HT). This was compared to the weight gain of children fed a STD infant formula (Formula 2). An exclusively breastmilk-fed group (BFD) served as a reference group for exploratory analysis.

#### Secondary objectives

To determine other measures of body composition (anthropometric data), the incidence of infections, digestive tolerance (flatulence, vomiting, and regurgitation), 3-day dietary daily intake, stool (consistency and frequency), behavior (restlessness, colic, night awakenings), the relative abundance of fecal bacterial species (fecal microbiota) and bacterial pathways (fecal metagenome), safety and tolerability of the infant formula at 2, 4, 6 and 12 months of life. Those outcomes were compared between groups fed formula 1 and 2 and each of them was compared with the BFD group.

The safety objectives included any adverse events arising from the consumption of the study formula.

### Trial design

The INNOVA study was designed as a randomized, multi-center, double-blind, parallel, and comparative clinical trial of equivalence of two starting infant formulas for infants. Furthermore, a third un-blinded group of breastfed infants was used as a further reference group for exploratory analysis. Following the current legislation of the EU on infant formulas (EC Regulation No. 1924/2006) for the demonstration of the nutritional and healthy properties of food intended for infants, it is not compulsory to carry out specific clinical tests. However, although this work is not a clinical drug trial, the ethical, methodological, and scientific rigor of clinical drug trials are included. The Gantt chart of this trial is shown in **Table 1**.

**Table 1.**
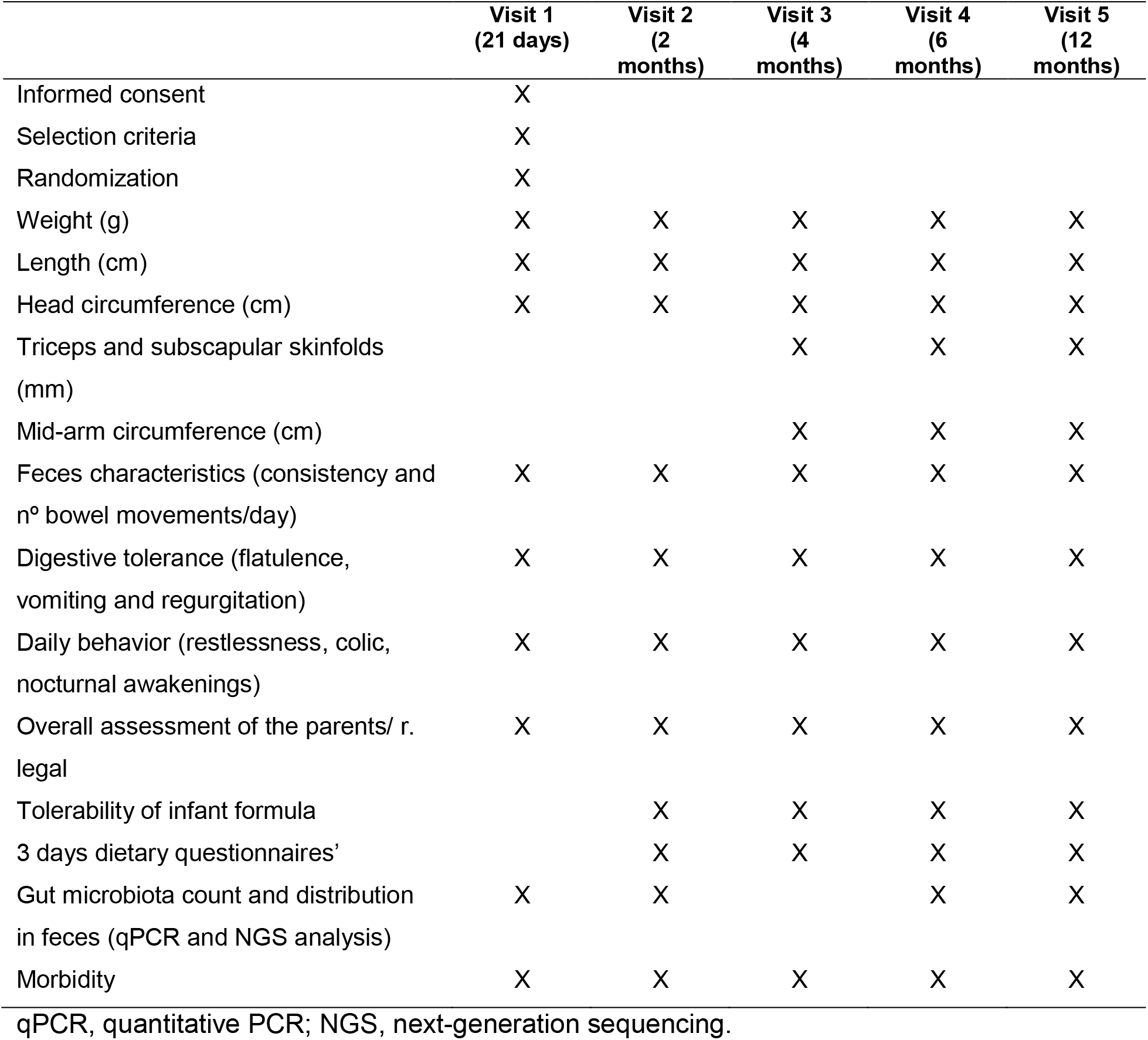
Gantt chart of the trial.

|  | Visit 1<br>(21 days) | Visit 2<br>(2 months) | Visit 3<br>(4 months) | Visit 4<br>(6 months) | Visit 5<br>(12 months) |
| --- | --- | --- | --- | --- | --- |
| Informed consent | X |  |  |  |  |
| Selection criteria | X |  |  |  |  |
| Randomization | X |  |  |  |  |
| Weight (g) | X | X | X | X | X |
| Length (cm) | X | X | X | X | X |
| Head circumference (cm) | X | X | X | X | X |
| Triceps and subscapular skinfolds (mm) |  |  | X | X | X |
| Mid-arm circumference (cm) |  |  | X | X | X |
| Feces characteristics (consistency and nº bowel movements/day) | X | X | X | X | X |
| Digestive tolerance (flatulence, vomiting and regurgitation) | X | X | X | X | X |
| Daily behavior (restlessness, colic, nocturnal awakenings) | X | X | X | X | X |
| Overall assessment of the parents/ r. legal | X | X | X | X | X |
| Tolerability of infant formula |  | X | X | X | X |
| 3 days dietary questionnaires' |  | X | X | X | X |
| Gut microbiota count and distribution in feces (qPCR and NGS analysis) | X | X |  | X | X |
| Morbidity | X | X | X | X | X |

qPCR, quantitative PCR; NGS, next-generation sequencing.

Parents were informed by pediatricians involved in the study of the possibilities of participating in it at a meeting at 15 days of infant age. If they accept to participate, they went a week later (visit at 21 days of age) to the health center for the baseline visit of the study. The pediatrician requested, at the meeting at 15 days of life, that parents provide at the next visit relevant information from the maternal history that was required for the study and that was not available in the pediatric history. The visit that took place within the scheduled time was considered valid with a margin of ± 3 days in the visit at 21 days of age, ± 1 week in the visit at 2 months of age, and ± 2 weeks in visits to 4, 6 and 12 months of age.

### Participants, interventions, and outcomes

#### Study setting

The present study was designed under the hypothesis that the weight gain achieved by infants fed formula 1 would be equivalent to that observed in children fed with formula 2 or STD. Secondarily, whether the formula 1 and 2 were equivalent in terms of weight gain in comparison with a breastfeeding group was also analyzed. To obtain greater robustness in the results obtained, a prospective, parallel-group, randomized, masked and comparative study was designed between two formulas of artificial feeding (formula 1 vs. formula 2) which, in turn, is compared with a third branch, not masked, of breastfeeding.

#### Eligibility criteria

Participants provided written informed consent before any physical assessments occurred.

### Inclusion criteria

Participating infants should meet all inclusion criteria in the absence of exclusion criteria. Inclusion criteria were: 1) Healthy children of both sexes; 2) Term children (between 37 and 42 weeks of gestation); 3) Birth weight between 2500 g-4500 g; 4) Single delivery; and 5) Mothers with a body mass index, before pregnancy, between 19 and 30 kg/m^2^.

### Exclusion Criteria

Volunteers were excluded from participation based on the following criteria: 1) Body weight less than the 5^th^ percentile for that gestational age; 2) Allergy to cow’s milk proteins and/or lactose; 3) History of antibiotic use during the 7 days before inclusion; 4) Congenital disease or malformation that can affect growth; 5) Diagnosis of disease or metabolic disorders; 6) Significant prenatal and/or severe postnatal disease before enrollment; 7) Minor parents (younger than 18 years old); 8) Newborn of a diabetic mother; 9) Newborn of a mother with drug dependence during pregnancy; 10) Newborn whose parents/caregivers cannot comply with procedures of the study; 11) Infants participating or have participated in another clinical trial since their birth.

### Recruitment strategy

In the trial, there was a preliminary information meeting in which the parents of 15-day-old children were informed about the nature of the study and the inclusion and exclusion criteria to participate in it.

Seven days later and in case of accepting to participate in the study, the initial visit was carried out with the signing of the informed consent and the randomization of the child to the treatment group. During the study, visits were made at the times 21 days, 2, 4, 6 and 12 months of age of the child, with ± 3 days for the visit at 21 days of age, ± 1 week for the visit at 2 months, and ± 2 weeks for the others. During the first 6 months of the trial and until obtaining the main variable, the child was maintained only with the starting formula or with natural breastfeeding in the case of group BFD. From 6 months, the same starting formula was maintained and complementary feeding (purées) were introduced with the usual foods according to the infant’s age (“*Guide for the introduction of complementary feeding*”, [25]). For the BFD group in the second 6 months of the study, a routine feeding was maintained, as if they would have not participated in the trial, namely, a normal complementary feeding was introduced.

### Trial variables

#### Main variable

The main efficacy variable was weight gain (g/day) between recruitment and 6 months of age (calculated as the difference in infant weight between the initial visit and the 6-month visit, divided by the number of days between both visits).

#### Secondary variables

- Weight and/or weight gain (g/day) at 21 days, 2, 4, 6 and 12 months of age.
- Length and/or length gain at 21 days, 2, 4, 6 and 12 months of age.
- Head circumference and/or head circumference gain at 21 days, 2, 4, 6 and 12 months of age.
- Triceps and subscapular skinfolds and/or gain of skinfolds at the times of 4, 6 and 12 months of age.
- Mid-arm circumference and/or mid-arm circumference gain at the times of 4, 6 and 12 months of age
- Characteristics of the feces (consistency and the number of stools/day) at times 21 days, 2, 4, 6 and 12 months of age.
- Digestive tolerance (flatulence, vomiting and regurgitation) at times 21 days, 2, 4, 6 and 12 months of age.
- Behavior (restlessness, colic, night awakenings) at times 21 days, 2, 4, 6 and 12 months of age.
- 3 days of dietary records using validated questionaries’ at times 2, 4, 6, 8, 10, and 12 months of age
- Counting, and identification of the gut microbiota in fecal samples (qPCR and bacterial DNA Next-Generation Sequencing, NGS), at 2, 6 and 12 months of age.
- Morbidity (within 1-year morbidity (within 1 year)).
- Tolerability of the products (in the course of 1 year).
- Global evaluation of the parents. Infant behavior Diary.

#### Complementary variables of the study

During all visits, parents were asked about digestive tolerance to the formulas (vomiting, gas, among others) as well as about the consistency of the stool and the behavior of the infant. This information is reflected in the Clinical Research Document (CRD). In addition, the parents had access, from an electronic device provided by the center, to a questionnaire where they could reflect on this type of event at the time it happened.

### Data quality

This included measures to ensure data accuracy and reliability including the selection of qualified investigators and site suitability, review of protocol procedures with the investigator and associated personnel before the trial, and regular monitoring visits by the sponsor. This was performed with the Contract Research Organization (CRO): Instituto Fundación Teófilo Hernando (IFTH) Parque Científico de Madrid. UAM. C/ Faraday 7. Edificio CLAID 28049. Madrid, Spain.

According to the Good Clinical Practice standards, the following were considered:

- Quality Control: “techniques and operational activities that are undertaken within the Quality Assurance system to verify that the quality requirements have been met in the activities related to the test.”
- Quality Guarantee: “all those planned and systematized activities that are established to ensure that the trial is carried out and that the data is generated, documented (recorded) and communicated following the Good Clinical Practice guidelines and the relevant regulations.”
- Source data “all the information contained in the original files and certified copies of the original files regarding clinical findings, observations or other activities of the clinical trial that are necessary for the reconstruction and evaluation of the trial.”
- The source data is contained in the source documents (the original files or the certified copies).
- Source documents: “original documents, data and records (e.g. clinical histories, clinical and administrative charts, laboratory reports, memoranda, subject diaries or assessment questionnaires, drug dispensing records, data recorded by computerized instruments, copies or transcripts certified after verification as true copies, microfiche, photographic negative, microfilm or magnetic media, X-rays, files of the subjects and records kept in the pharmacy, in the laboratories and the medical-technical departments involved in the clinical trial).

Monitoring is the “act of supervising the development of a clinical trial, and ensuring that it is carried out, filed and published following the protocol, the Standard Operating Procedures (SOP), the standards of the GCP guide, as well as current regulations. Therefore, the person responsible for monitoring was responsible for verifying that:

- The rights and welfare of human subjects are protected.
- The data obtained in the study are accurate, complete, and verifiable with the source documents (in the case of this study, the clinical history).
- The conduct of the study is following the protocol and the approved amendments, with the guidance of GCP, and with current legislation.

The person in charge of monitoring followed his own monitoring guidelines and SOPs and made the monitoring visits to the center as many times as necessary, following the monitoring plan agreed upon with the sponsor. Likewise, the study monitor was independent of the research team.

### Deviations or violations of the protocol

Any non-compliance with the assay protocol or GCP is considered a protocol violation or deviation. In case of detecting any deviation, the corresponding corrective measures were defined and applied as soon as possible. However, no deviation from the protocol was applied whether it has not been previously reviewed and approved by “*Alter Farmacia, S*.*A*.” and by the Drug Research Ethics Committee, unless this was necessary to avoid an immediate risk to the study participants. In this case, both were notified of the application of the deviation as soon as possible. In addition, deviations and violations of the protocol were documented in the CRO deviation register.

### Limitations of the design

The main limitation is the foreseeable difficulty of recruitment and the permanence throughout the study both in the formula-feeding groups and in the case of breastfeeding. The group of BFD is clearly different from those artificially fed groups: differences in the lesson derived from maternal preferences, the impossibility of double-blind administration and heterogeneity due to non-randomization for all of which its comparison with breastfeeding artificial remains as an exploratory analysis.

### Criteria for premature withdrawal, termination and suspension of test

Participants had the right to withdraw from the study at any time and for any reason, without giving explanations or suffering any penalty for it. Likewise, the investigator could withdraw study participants if they did not comply with the study procedures or for any reason that, in the investigator’s opinion, may pose the infants at risk or makes it necessary to suspend breastfeeding. If a child was diagnosed during the trial with any disease that required prolonged antibiotic treatment (more than 10 days) or required changes in the breastfeeding/nutrition pattern was also withdrawn from it. Infants who were withdrawn or dropped out of the trial were followed up by their pediatrician as they would be in routine clinical practice.

In the event of any withdrawal, the circumstances must be detailed on the CRO study completion page. Likewise, the data of the subjects who dropped out or were withdrawn from the study were also documented in the final report of the study’s clinical trial. The trial was considered finished when: all the participating infants had finished their last visit and the monitor during his visits had verified that the trial had been carried out, filed, and published following the protocol, SOP and the GCP guidelines, as well as current regulations. On the other hand, the trial could be suspended, interrupted or terminated prematurely by the trial promoter at any time with the corresponding communications and justification to all parties involved.

### Sample size

The sample size of the trial is 210 children (70/group), based on the main outcome of weight gain, which is the main variable chosen according to the guidelines “Guidelines from the American Academy of Pediatrics Task Force on Clinical Testing of Infant Formulas” [26]. The infants were selected by Primary Care pediatricians in competitive recruitment. The study was carried out in 21 centers, all located in Spain, of which 17 recruited at least one subject. In total, 217 subjects signed the informed consent (IC) and 145 were randomized to receive one of the two infant formulas. Of these 145, 3 were randomization failures and 2 were screening failures, thus 140 infants who met all the inclusion criteria and no exclusion criteria were included in the study, and 70 were unblinded within infants exclusively breastfed. A total of 185 subjects completed all study visits, of which there were 25 dropouts, 12 in the breastfeeding group, 8 in the formula 1 group, and 5 in the standard formula group.

**Figure 1.**
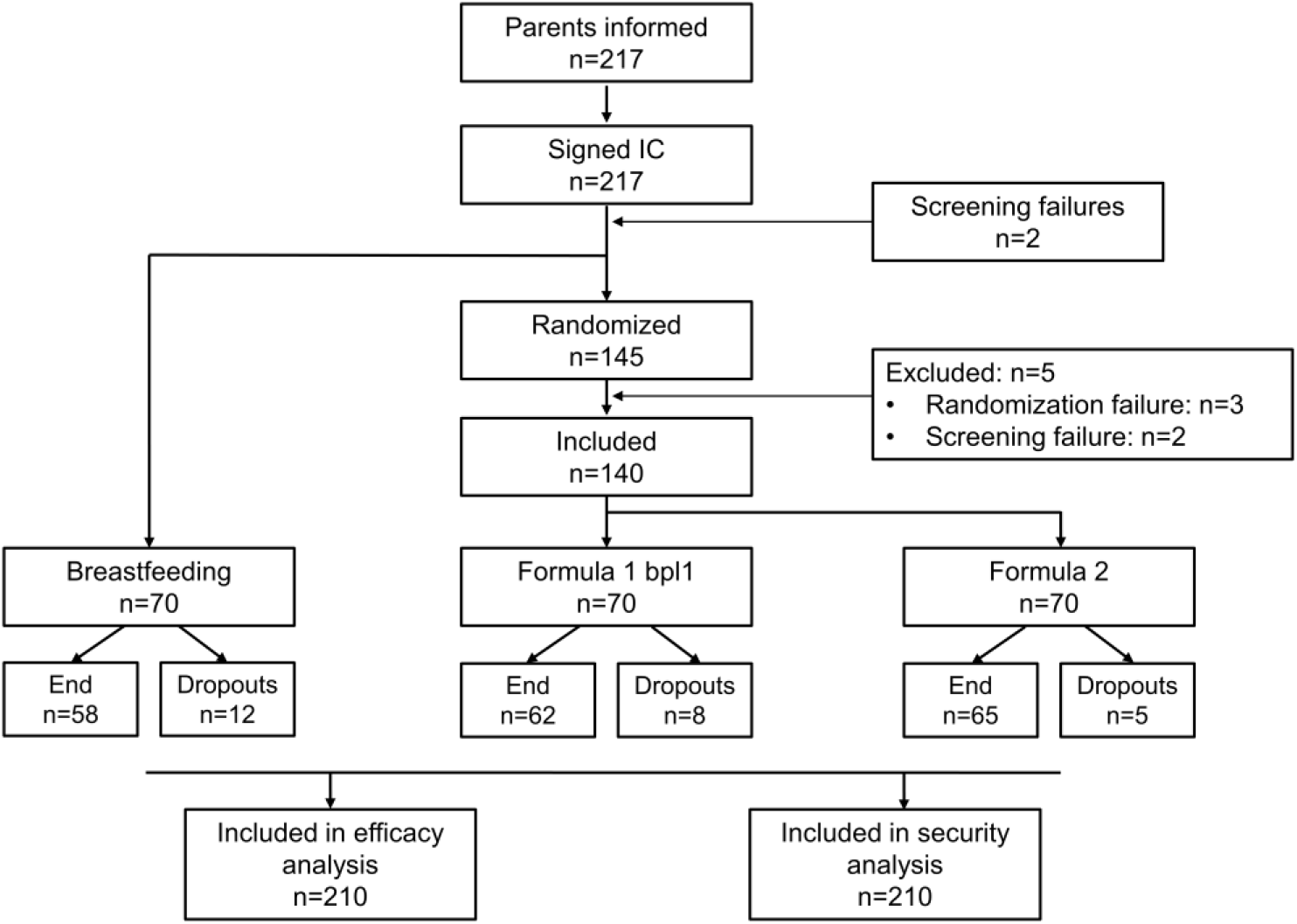
summarizes the disposition of the subjects in the study. IC, informed consent.

### Assignment of interventions

#### Managed Products

- Group 1 (Formula 1): Nutribén Innova® 1
- Group 2 (Formula 2): Nutribén® Standard
- Group 3: Breastfeeding (External control exploratory analysis)

Infants were recruited from Primary Care Pediatrics clinics by the pediatricians participating in the trial. Each pediatrician informed and proposed to the parents of 15-day-old infants who come to their office routinely (for regular medical check-ups) to participate in the trial. Infants that were not a candidate for breastfeeding (for different reasons) were proposed to participate in the formula-feeding groups. To keep the three arms of the trial balanced, one candidate breastfeeding subject was recruited at each center for every two infants fed with infant formula. The selection of the children of the BFD group was made among those infants who met the inclusion and exclusion criteria of the study and fit the categories of sex, BMI of the mother before pregnancy (<25, >25kg/m^2^), and birth weight (<3500 g, >3500 g) of the last infant in the formula-fed group included in the center.

#### Identity of the products

The experimental product object of this trial INN (Formula 1 or Nutribén Innova^®^ 1) and the STD formula (Nutriben^®^) comply with the recommendations of the ESPGHAN (European Society of Pediatric Gastroenterology, Hepatology and Nutrition) and with Regulation 609/2013 of the European Parliament and of the Council regarding foods intended for children, infants and young children, foods for special medical purposes and complete diet substitutes for weight control and repealing Council Directives 92/51, Directives 96/8/EC, 1999/21 /CE,2006/125/CE and 2006/141/CE of the Commission, Directive 2009/38/CE of the European Parliament and of the Council and Regulations 41/2009 and 953/2009 of the Commission. The following table summarizes the main characteristics of the study formulas (**Table 2**). More detailed information on the composition of each of the products can be found in Additional information (Supplementary Table S1).

**Table 2.** Comparison of the composition of the study formulas

|  | **Formula 1 (Nutribén Innova®1 or INN)** | **Formula 2 (Nutriben® or STD)** |
| --- | --- | --- |
| *Fat | Vegetable oils and fat milk | Vegetable oils |
| *GOS | Yes | Yes |
| Proteins | 8% less protein per 100 kcal than the standard. 70:30 Whey protein: Casein enriched with α-lactalbumin (20 % of total proteins) | 60:40 whey protein: casein. α-lactalbumin (10 % of total proteins) |
| DHA/ARA | -AA 24 mg/100 kcal<br>-DHA 24 mg/100 kcal | -AA 10 mg/100 kcal<br>-DHA 10 mg/100 kcal |
| BPL1™ HT | Yes (thermally inactivated) | No |

\*Same amount per 100 kcal for both formulas
GOS=galacto-oligosaccharides; DHA=docosahexaenoic acid; ARA=arachidonic acid; BPL1™ HT = *Bifidobacterium animalis* subsp. *lactis*

### Packaging and labeling

Blinding for both investigator and participant remained assured as both infant formulas were labeled the same. The study’s powdered infant formulas were packaged in 800 g net weight cans. They included, among other aspects, the following information:

- Lot number
- Expiration date.
- Net weight (800g)
- Preparation, use and maintenance instructions.
- Name and address of the manufacturer.
- List of Ingredients
- Nutrients
- Identification code of the formula.

### Administration of the product to the affiliated centers

The promoter supplied each center participating in the study with a sufficient quantity of product. The researchers were in charge of calling the promoter to request the formulas as they run out. Each investigator should record the amount of investigational product dispensed by the promoter and the amount returned with their corresponding dates. Likewise, the CRD of each subject should reflect the quantity of product delivered and the date, as well as such as the quantity of product or returned and its date. During visits to the center, the monitor verified the amount of investigational products dispensed, returned, and stored in the center. At the end of the trial, all the remaining product was delivered to the promoter to carry out the necessary analyses and finally discarded.

### Administration route

Infant formulas were administered *ad libitum* orally. The two trial formulations were administered following the preparation instructions in the manufacturer’s package insert.

### Agreement

The parents or legal representatives filled in the intakes of the corresponding infant formula and its quantity in the 3-day dietary diaries. In addition, at each visit, they must return to the center all the containers that they have leftover (empty containers and with the product). The objective is to record the amount of product dispensed to the participants and the amount of infant formula that has been used up to the time of visit. These registries took place from the second visit to the study. Upon completion of the study (at the 12-month visit), parents or guardians should return all bottles of infant formula to the health center. Participants in this study were considered compliant if they had taken at least 80% of the formula during the first six months. In the case of the BFD group, infants should exclusively consume breast milk on demand. A valid external control was considered if 80% of the feeds or more have been from breastfeeding. From the sixth month of life, it is necessary to introduce complementary feeding since breast milk or infant formulas by themselves are no longer sufficient to guarantee the correct growth and development of infants. However, dairy consumption after six months is still necessary. The most widespread recommendation establishes that dairy consumption during the first year of life should be maintained at around 500 mL per day. Likewise, it is important to point out that it is not advisable to increase milk consumption above this amount because it would displace the consumption of other types of complementary foods and, in addition, it seems to hinder the ability to eat.

### Randomization, masking and assignment of the formula

Study subjects were randomly assigned to either group 1 of the Nutribén Innova® 1 study (Formula 1) or group 2 of the Nutribén® Control (Formula 2). The assignment of each participant was carried out by randomization in blocks balanced by a center and stratified by sex of the infant, BMI of the mother before the pregnancy (<25, ≥ 25 kg/m^2,^ and birth weight (<3500 g, ≥ 3500 g) using the “block.random” function of the R software, to maintain a parallel design. If it was necessary to unmask the randomization, a representative of the center, in this case, the researcher had to request authorization from a person from the Promoter, thus that through the CRD he can know the formula assigned to that subject in question.

### Medications or treatments not allowed during the trial

The following treatments were forbidden in the trial:

- Antibiotics during the 7 days before inclusion.
- Antibiotic for 10 days during the trial.
- Any treatment, food or product that at any time during the trial can interfere with it
- Breastfeeding different from that of the group assigned in the trial.

Any treatment, food or product that is going to be administered to the infant must be previously consulted with the research team of the trial. Any concomitant treatment during the trial will be recorded in the CRD.

#### Allocation

The trial was expected to last approximately 2 years, consisting of the following stages:

- Commissioning 4 months.
- Start-end of recruitment: 12 months.
- Experimental-clinical phase: 12 months
- Analysis of results and final report: 3 months

The total duration of participation in the trial for each infant is until the infant turns one year old (because if he completes the entire trial, the last visit is at 12 months).

#### Data collection

The data for this trial were obtained from original documents and records of each participant related to the trial such as the CRD and the database. For this essay, source documents are specifically considered: the clinical history reports, laboratory reports, evaluation diaries or questionnaires, and files of the participating subjects.

#### Data management and archiving of records

All study data were recorded in physical documents (Clinical History) and computerized CRD. The data collected in the CRD of each infant included in the essay is transferred to a computerized file (database) that allows its processing using a statistical analysis program. This codified information lacks personal data of the participant’s object of this essay, so it is not registered with the Spanish Agency for Data Protection.

#### Electronic data collection notebook and confidentiality

In this trial, a CRD was reinforced to enter the study data. The investigators were responsible for ensuring the integrity and accuracy of the CRD. A CRD was completed for each subject participating in the study. If erroneous data is entered and sent, this error should be reported as soon as possible to the person in charge of managing said data. This CRD was specifically designed for this study. In addition, the system automatically performed an audit trail of all entries and corrections made in the CRD. Only authorized personnel had access to it through the combination of an identification code and a password. By entering these keys, one could enter, modify or view study data. The information of the participants was treated in an encrypted manner. Subjects were identified by a code made up of nine characters. Thus, the content of the CRD, as well as the database where the information was recorded. They were encrypted and protected from uses not allowed by people outside the company, therefore, were considered strictly confidential. The treatment, communication and transfer of personal data of all participating subjects complied with the provisions of current legislation related to personal data protection.

### Measurements and evaluations

#### Anthropometric measures

Weight, length, and head circumference measurements were taken at all study visits. From the visit scheduled for the fourth month of life, the mean arm circumference, the triceps skinfold, and the subscapular skinfold were added to the previous measurements. All these measurements were done in duplicate. Using as valid data the mean between the two. Each center had all the necessary materials to carry out these measurements. They needed to always use the same material with all subjects and, of course, the material should be calibrated.

#### 3-day dietary questionnaires’

The dietary record or diary is one of the available methods for obtaining data on dietary intake. To be representative, it must be carried out for at least three days. This diary makes it possible to record the subject’s daily food consumption, allowing to know the information about the type and amount of food, the way of preparation and the hours in which the meals are made. For this reason, in this trial, parents or guardians were asked to complete the 3-day dietary diary for visits 2, 3, 4, and 5, and two additional diaries at 8 and 10 months of age, to know the diet of their infants.

#### Introduction of Complementary Feeding

Complementary feeding was implemented in a specific way and controlled by the pediatrician according to the “Guide for the introduction of complementary feeding”, the food introduction scheme that was carried out throughout the study is described. This scheme was developed using as a basis the “Guide for the introduction of complementary feeding” from the “Hospital Infantil Universitario Niño Jesus”, Madrid, Spain, In addition, if parents or guardians choose to give commercial infant food, they were offered the option of providing them with some of the Nutribén ® brand children’s products free of charge.

#### Fecal samples

Fecal samples were collected at 21 days, 2, 6 and 12 months of infant life. To do this, the center provided the parents and/or guardians with all the necessary materials for collection. Center staff labeled the sample so that the patient identifier, the date the sample is collected, and the date the sample was shipped appeared. The parents and/or guardians should store refrigerated (24 h maximum) and later store in the freezer at -20°C±1°C. For the transport, samples were maintained on dry ice to keep the optimal conditions for their subsequent analysis. These samples arrived at the laboratory in a maximum of 10 hours and, once received, temperature control was assessed. On the other hand, to guarantee the viability of all the samples, no more than six months passed from the time they were collected at the center until they arrived at the laboratory. The analysis of gut microbiota, short-chain fatty acids (SCFAs) and Immunoglobulin A (IgA) was carried out by the company “Biópolis S.L” following standard procedures.

##### Sequencing procedure

An analysis of the gut microbiota was carried out from the fecal samples to determine differences over time and between groups. DNA extraction was carried out using an optimized protocol, which includes a combination of bead beating and enzymatic lysis, using a modified protocol [27], and applying the QIAmp Power Fecal Kit (Qiagen). DNA quality control was carried out by using the Nanodrop equipment (ThermoFisher). Subsequently, a library was generated per sample of PCR products from the amplification of the hypervariable region between V3-V4 of the bacterial 16s rRNA [28], marked with a molecular identifier (ID) and performed a primer dimer cleanup. The libraries were sequenced on the Novaseq platform of Illumina in combination of 250PE. A negative control containing water was used to confirm the absence of contamination. For the annotation of the sequences obtained in the sequencing process, a series of quality control steps were previously carried out. The first step was focused on cleaning the sequences. Due to the sequencing method, two sets of sequences were obtained for each of the samples, one file with the sequences sequenced in one direction (R1), and another file sequenced in the other direction (R2). Each pair of sequences is subjected to overlap between them, to obtain a unique and complete sequence, using the PEAR V.0.9.1 program [29] (http://www.exelixislab.org/web/software/pear) with the default parameters except the minimum overlap parameter between the sequences at each end, which was set at 70nts, obtaining at the end a single file with the total number of overlapping sequences. Once the overlapping sequences were obtained, they were processed by quality to eliminate those sequences with low quality. First, the ends of sequences that did not reach the quality of Q20 were eliminated. Once the poor quality ends were eliminated, the sequences whose average quality value was less than Q20 were eliminated. The remaining sequences were used in the following steps. With the quality sequences, residues of amplification primers were eliminated, with which it was possible to reduce the bias that can be introduced in the process of amplification. Once the primers had been removed, sequences longer than 200nt in the 16S sequences were selected since the smaller sequences have greater probability of being erroneously associated with taxonomic groups. These two steps were performed with the CUTADAPT program version 1.8.1 [30]. The last step of quality processing was the removal of chimeric sequences resulting from amplification. This step is performed with a previously designed database and with the Uchime program [31]. Thus, the sequences that were later used for the annotation process were obtained. To reduce the complexity of the annotation process, sequences sharing 97% similarity were grouped into a single sequence using the “cd-hit” program to that only those sequences that were significant were annotated, and the results were applied to the group of sequences represented by the analysis. Sequence clusters were compared against an rRNA database using the local alignment BLAST strategy to associate each cluster with one of the database’s taxonomic groups.

All statistical analyses were performed using the R programming language. The diversity of the samples was studied using the *vegan l*ibrary. On the one hand, alpha diversity indices such as Shannon, Simpson and richness index were studied. On the other hand, beta diversity was also studied using Bray-Curtis distances and multidimensional ordering techniques such as PCoA and Non-Metric Multidimensional Scaling (NMDS). PERMANOVA tests were performed to verify the significance of these results. A comparative study was carried out between the 4 times of the study and the 3 proposed treatments. This study was elaborated using the DESeq2 tool, in which it is assumed a negative binomial distribution in the matrix of counts to proceed with the statistical tests of Wald that allow discerning if there is a differential effect according to the time or treatment between the samples.

##### Analysis of short-chain fatty acids in feces

SCFAs were measured in feces since it has been observed that they produce effects on lipid metabolism and adipose tissue at different levels. High-performance liquid chromatography (HPLC) was used to quantify the amount of SCFA (acetic acid, butyric acid, lactic acid, propionic acid and succinic acid) in feces. The samples were separated, filtered and processed. SCFA, lactic acid and sulfuric acid solution were purchased from MilliporeSigma (Burlington, MA, USA). Also, the Aminex HPX-87H column (300×7.8mm, 9um particle size) and Cation H micro-guard cartridge (30×4.6mm) were purchased from Bio-Rad Laboratories (Hercules, CA, USA). The separation system consisted of an Alliance 2695 HPLC equipment (Waters Corp., Milford, MA, USA) equipped with a refractive index detector (RID 2414, Waters) and the computer software Empower 3.0 (v. 7.4). An ion-moderated partition chromatography column (Aminex HPX-87H with Cation H micro-guard cartridges) was used. The column temperature was maintained at 60ºC, and the different organic acids were eluted with 5mM H_2_SO_4_ prepared using Milli-Q filtered water at a flow rate of 0,6ml/min. The RID internal temperature was 45ºC and the injection volume 20 μl. Sample preparation includes a 1:4 aqueous dilution and filtration through 0.22um PVDF filters. Peaks were identified by comparison to retention times of analytical standards and quantified using standard curves of the different SCFA (acetic, butyric and propionic acids) and other organic acids (such us lactic acid) [32].

##### Calprotectin analysis (ELISA)

The evaluation of calprotectin in feces is a reliable method, which enables the distinction between organic diseases and functional gastrointestinal diseases [33]. Calprotectin was determined in fecal samples by using a ELISA kit (BÜHLMANN fCAL).

##### Secretory Immunoglobulin A Assay analysis (ELISA)

The levels of fecal secretory IgA (sIgA) are a good representation of that found in the colon [34]. The importance of fecal sIgA determination lies in the fact that it plays an important role in the defense of the gastrointestinal tract since, the greater the amount of sIgA secreted in feces, the greater the antiviral and antimicrobial capacity the subject present. The amount of sIgA in feces was determined by the ELISA technique (IDK® sIgA ELISA, Immundiagnostik AG, Bensheim, Germany).

#### Statistical methods

The main objective of the study was to evaluate whether the weight gain was equivalent between treatment groups receiving formula 1 and 2, and it has been decided that the sample size of the trial is 210 children (70/group). Previous studies carried out in children fed from 0 to 6 months with different formulations of infant formula have shown a mean weight gain of around 20-25 g/day with a standard deviation between 5 and 6 g/day. In most of these studies, a difference in mean weight gain of 3 g/day was considered clinically relevant. Thus, the main objective will be resolved by using a t-test for independent samples. Considering a power of 80%, a significance level of 5%, an equivalence limit of 3 g/day, and a common standard deviation of 5.5 g/day, we would need to recruit 59 children in each of the groups. If we consider a loss rate of 20%, it will be necessary to include 70 children in each of the groups, which means 140 infants. A third group of the same size (70 children) should be included for the secondary comparisons between the bottle-fed and BFD groups, maintaining the same significance and power in this secondary comparison as in the main comparison. Categorical variables will be described as frequencies or percentages. Continuous variables will be described as mean and standard deviation. Between-group differences in measures of growth and body composition will be analyzed using analysis of variance, including analysis of covariance and analysis of variance for repeated measures when required. The Chi-square test will be used to compare discrete variables between groups. The Bonferroni correction will be used when comparisons are made between more than two groups. The alpha level of significance is set at 0.05. All evaluable infants will be included in the statistical analysis, considering “evaluable” all those who have the main measurement variable. Statistical analysis will be performed both by intention to treat and by protocol.

## Ethics and dissemination

### Research ethics approval

This clinical trial was carried out following the recommendations of the International Conference on Harmonization Tripartite on PCBs, the ethical-legal principles established in the latest revision of the Declaration of Helsinki, as well as the current regional regulations that regulate pharmacovigilance and food safety.

### Drug Research Ethics Committee approval

This protocol and any material that is delivered to the subject (information documents for the participant, informed consent, etc.) must be delivered to the Ethics Committee of the Drug research (ECDR) as well as any advertising material or compensation offered to the participating subjects. Before starting the study, the approval of the ECDR was obtained. This approval was documented in a letter sent to each researcher specifying the date on which the ECDR met to give its approval.

### Benefit-risk assessment for the research subjects

The infants included in the trial receive BFD or the formula they need according to the preferences and possibilities of the mother (breastfeeding or one of the two artificial formulas at the start of the trial). Both formulas comply with the legal requirements applicable to infant formulas. Infants assigned to one of the two formula-feeding groups in the trial received the corresponding formula free of charge. On the other hand, the parents of the BFD group received financial compensation for their participation in the test. During the trial, the appearance of adverse reactions was monitored and, if they appear, they were communicated in a pertinent manner.

### Consent or assent

Before inclusion in the trial, the minor’s legal representative was duly informed, providing an information sheet together with the corresponding Informed Consent. The rationale and objective of the trial were explained, with the risks and benefits that may arise, as well as the procedure to follow. Both documents are attached as Annexes to this protocol.

### Confidentiality

The information of the participants was treated in an encrypted manner. Subjects were identified by a code made up of nine characters. The first two digits identified the center, the next three e the patient number, and the last four corresponded to the randomization code. This code was used to identify the subjects in the CDRe and the rest of the documents presented to the promoter. The content of the CRDe, as well as the database where the information was recorded, was encrypted and protected from uses not permitted by people outside the research and, therefore, were considered strictly confidential. The treatment, communication and transfer of the personal data of all the participating subjects were adjusted to the provisions of current legislation related to the Protection of Personal Data. All personal data obtained in this study are confidential and were treated following the Spanish Organic Law 3/2018, of December 5, on the Protection of Personal Data and guarantee of digital rights. The researcher or the institution allowed direct access to the data or source documents for monitoring, auditing, and review by the ECDR, as well as inspection of the trial by the health authorities.

### Access to data

The study coordinator and principal investigators will have access to the cleaned datasets. Principal investigators will oversee intra-study data sharing. To ensure confidentiality, data shared amongst other study investigators will be identified.

### Ancillary and post-trial care

We expected this trial would have minimal risks to participants. However, if a serious or unexpected adverse event related to trial procedures occurred, participants would have provisional care beyond that immediately required.

### Dissemination policy

Data from this trial will not be presented in public or submitted for publication without the permission of the principal investigators. With the permission of principal investigators, the results of this trial will be disseminated via scientific symposia and conferences and will be published in peer-review scientific journals. The requirements for authorship will adhere to scientific journal guidelines.

## Discussion

The WHO recommends exclusive breastfeeding for 6 months as the best way of feeding infants, where they obtain only breast milk with no other liquids or solids, except for oral rehydration solution, drops/syrups of vitamins, minerals or medicines [35]. Despite the WHO guidelines, currently, the rates of exclusive breastfeeding for the first 6 months remain low. Generally, there are several obstacles such as maternal smoking, educational level or the need to return to work accounting for the low rates of exclusive breastfeeding [36-38].

In those cases where breastfeeding is not possible, it is necessary to have infant milk formulas that at least promote proper growth and development. Even though efforts have been taken to partly mimic the nutrition profile of human breast milk in the infant formulas for normal infant growth and development, there is a need for further studies to improve it as infant formula is not without limitations.

The present trial is a randomized double placebo-controlled clinical trial designed to provide evidence regarding the beneficial health effects of a novel starting infant formula with a lower amount of protein and the proportion of casein to whey protein ratio of about 70/30 by increasing the content of α-lactalbumin, higher levels of DHA and ARA, and containing a post-biotic thermally inactivated (BPL1™ HT), compared with a standard infant formula. An exclusively breastfed population was followed-up as a reference.

Besides the metabolic and endocrine exposures during the pregnancy [39], the protein intake in formula-fed infants during the first year of life impacts on growth later obesity risk and associated metabolic disorders in adulthood [15, 40]. In fact, the protein content of an infant formula is usually higher than that of human milk to provide all essential amino acids in adequate amounts [41]. Nonetheless, infants fed a formula containing higher protein gain more weight during the first year of life and are heavier at 2 years of age than are infants fed a lower-protein formula [40], which reduces BMI and obesity risk at the school stage (at 6 years of age) [42]. Here, the experimental formula (Nutribén Innova^®^) contains 8 % less protein per 100 kcal than the standard one, being within the range recommended by the EFSA after proposing to reduce the maximum protein content of infant formulas to 2.5 g/100 kcal. In addition to the total protein content, the whey/casein ratio in infant formulas is crucial for the first year of life. Whey proteins, particularly β-lactoglobulin and α-lactalbumin, are rapidly digested and participate in the building of muscle mass, as they remain mostly soluble in the stomach and pass more rapidly toward the intestine [43]. Proteins in human milk are present in two major compartments, whey and casein micelles, as well as other, but minor compartments, such as cells, milk fat globule membranes and exosomes [44]. Casein proteins are water-insoluble high molecular weight molecules mainly made up of three casein subunits: α-S1-casein, β-casein, and κ-casein [45]. The main biological function of these fractions is the primary source of phosphate and calcium in human milk, because of the highly phosphorylated nature of β-casein and α-S1-casein, and the requirement for calcium in forming the aggregates of casein micelles [46]. However, the amount should be controlled to avoid improper digestion in early life. Regarding the whey fraction, α -lactalbumin is the major protein in human milk, and the addition of bovine α-lactalbumin to infant formula modifies the plasma amino acid pattern of the receiver infant, allowing a reduction in the protein content of the formula. In this context, using cow’s milk as the protein source in infant formula might result in an α-casein-dominant formula, which differs from the whey protein dominance in human milk. Besides, bovine whey contains a high concentration of β-lactoglobulin, which is absent in human milk [47], therefore, adding bovine α-lactalbumin to infant formula has been suggested, because it modifies the plasma amino acid pattern in infants, making it more similar to that of breastfed infants [48]. Moreover, α-lactalbumin is relatively rich in tryptophan, which may result in satisfactory growth and plasma tryptophan levels similar to those of breast-fed infants and infants fed standard formula [49]. The experimental formula contained a reduction in α-casein content (70:30 whey: casein ratio vs 60:40 in the standard formula), and was enriched with α-lactalbumin, containing the 20 % of total proteins, compared to the 10 % of α-lactalbumin contained in the standard formula.

Regarding the lipid profile content, the experimental formula contained an equal amount of DHA and AA, 24 mg of each one per 100 kcal compared to 10 mg of each fatty acid per 100 kcal in the standard formula. Maintaining or improving the proportion of fatty acids present in infant formulas is equally important to ensure the correct growth and development of infants. Thus, in the last 20 years, formulas have been supplemented with LC-PUFA in amounts similar to human milk. Despite the new EU regulation that indicates that AA is not needed to be included, intervention studies assessing the impact of DHA- and AA-supplemented formulas have resulted in numerous positive developmental outcomes (closer to breast-fed infants) including measures of specific cognition functions, visual acuity, and immune responses [50-52]. AA has different biological functions compared to DHA, for example, AA has exclusive functions in the vasculature and specific aspects of immunity. Undeniably, most of the trials include both DHA and AA, and test development specific to DHA such as neural and visual development. DHA suppresses membrane AA concentrations and its function. Infant formula with DHA and no AA runs the risk of cardio and cerebrovascular morbidity and even mortality through suppression of the favorable oxylipin derivatives of AA [53]. Indeed, a trial contrasted the effects of four formulations in the long-term dose-response effects of LC-PUFA-supplemented formula feeding during infancy: 0.00% docosahexaenoic acid (DHA)/0.00% arachidonic acid (AA), 0.32% DHA/0.64% AA, 0.64% DHA/0.64% AA, and 0.96% DHA/0.64% AA against a control condition (0.00% DHA/0.00% AA). That study revealed an improvement in cognitive outcomes for infants fed supplemented formulas. However, it showed a reduction of benefit for the highest DHA dose, but even more importantly, AA levels exhibited a strong inverted-U function in response to increased DHA supplementation, implicating that the DHA/AA balance is vital in the contribution to cognitive and behavioral development of infants [54]. Hence, international expert consensuses recommend the use of an infant formula that provides DHA at levels of 0.3% to 0.5% by weight of total fat content and with a minimum level of AA equivalent to the DHA content, and should be clinically tested [18, 19]. Here, we will provide evidence about weight gain and body composition, among other outcomes, by testing an advanced formula enriched with a double amount of DHA/AA compared with a standard formula.

The experimental formula was also supplemented with a thermally inactivated postbiotic *Bifidobacterium animalis* subsp. *lactis*, BPL1™, which might confer some benefits concerning body composition, metabolism, and gut microbiota composition. Probiotics have numerous health benefits by modulating the gut microbiome; however, techno-functional limitations have made it gradually shift from viable probiotic bacteria towards non-viable postbiotics, paraprobiotics and/or probiotics-derived biomolecules, so-called postbiotics [55]. Postbiotics are emerging concepts and are not well defined yet, and indeed, other related terms have also been used, including ‘parapsychobiotics’, ‘ghost probiotics’, ‘metabiotics’, ‘tyndallized probiotics’ and ‘bacterial lysates’ [56] However, the field would benefit from merging around the use of a single, well-defined and understood term rather than the use of disparate terms for similar concepts. Here, we use the term ‘postbiotic’ to refer to BPL1™ HT, as the International Scientific Association for Probiotics and Prebiotics (ISAPP) reviewed and accepted this term in 2019 as useful for scientists, clinical triallists, industry, regulators, and consumers. The panel defined a postbiotic as a “preparation of inanimate microorganisms and/or their components that confers a health benefit on the host” [57]. The probiotic strain *Bifidobacterium animalis* subsp. *lactis*, BPL1™, reduces total lipid and triacylglycerols in the nematode *C. elegans*, increasing the survival, and modulating the tryptophan metabolism [58]. Moreover, BPL1™ decreases food intake, plasma ghrelin levels, decreased plasma tumor necrosis factor α and plasma malondialdehyde concentrations as a biomarker of oxidative stress in obese rats compared to a control group. Also, the postbiotic i.e inactivated BPL1™, reduced the ratio of plasma cholesterol total/LDL-cholesterol in obese rats, indicating an improvement in cardiovascular risk [22]. On the other hand, inactivated BPL1™, reduced BMI, waist circumference and visceral fat in individuals with abdominal obesity after twelve-week treatment [23]. Since the experimental formula of the present study was supplemented with BPL1™ HT, it should be noted that in vivo studies reported that this strain can be considered a Generally Recognized as Safe (GRAS) substance [26]. In the present trial, we will analyze the gut microbiota composition and functional pathway profiles in the infants fed with either formula 1 or 2, compared with the breastfeeding group as reference. This will provide solid evidence for the supplementation of BPL1™ HT in infant formulas.

In conclusion, this is a randomized, multicenter, double-blind, parallel and comparative clinical trial involving the evaluation of a novel infant formula with reduced content of total protein and modification of the whey/casein ratio of about 70/30 by increasing the content of α-lactalbumin, increased levels of both AA and DHA, and containing the postbiotic BPL1™ HT), in comparison with standard infant formula. A third unblinded group of breastfed infants was used as an additional control group for exploratory analysis. The safety, tolerance, and effects on growth and incidence of major acute infectious diseases and changes in gut microbiota are evaluated for 6 months, and until 12 months after the introduction of complementary feeding. This study will provide dense evidence regarding the beneficial health effects of having a novel starting infant formula over infants fed standard formula.

## Data Availability

All data produced in the present study are available upon reasonable request to the authors

## Additional information

Supplementary table S1.

## Acknowledgments

We thank to the pediatrician team who participated in this study: Alba Corrales; Alfonso Carmona; Amalia López; Ana Isabel Rodriguez; Ana Maderuelo; Carlos Nuñez de Prado; Eduardo Ortega; Esther Martin; Eva Castillo; Isabel Mayordomo; José Fernando Ferreira; José María Aguilar Diosdado; Maite Santos; Mari Carmen Pino Zambrano; María José Carnicero; Maria Teresa Santos; Vasilica Doina and Xavier Riopedre. We also thank to Ana Terrén-Lora, member of IFTH for her contribution to this study.

## Availability of data and materials

The datasets used and/or analyzed during the current study are available from the corresponding author upon reasonable request.

## Ethics approval and consent to participate

Ethics approval has been approved by “*Alter Farmacia, S*.*A*.” and by the Ethics Committee of the Drug research (ECDR). Written informed consent were obtained from all participants.

## Consent for publication

Not applicable.

## Author details

AGG, IG and MM were responsible for a careful review of the protocol, design, and methodology. FJRO, JPD and AG provided continuous scientific advice for the study and the interpretation of the results. These authors also wrote and critically reviewed the manuscript. All authors approved the final version of the manuscript. We thank to the pediatrician team who participated in this study: Alba Corrales; Alfonso Carmona; Amalia López; Ana Isabel Rodriguez; Ana Maderuelo; Carlos Nuñez de Prado; Eduardo Ortega; Esther Martin; Eva Castillo; Isabel Mayordomo; José Fernando Ferreira; José María Aguilar Diosdado; Maite Santos; Mari Carmen Pino Zambrano; María José Carnicero; Maria Teresa Santos; Vasilica Doina and Xavier Riopedre. We also thank to Ana Terrén-Lora, member of IFTH for her contribution to this study.

---

**Supplementary table S1.**
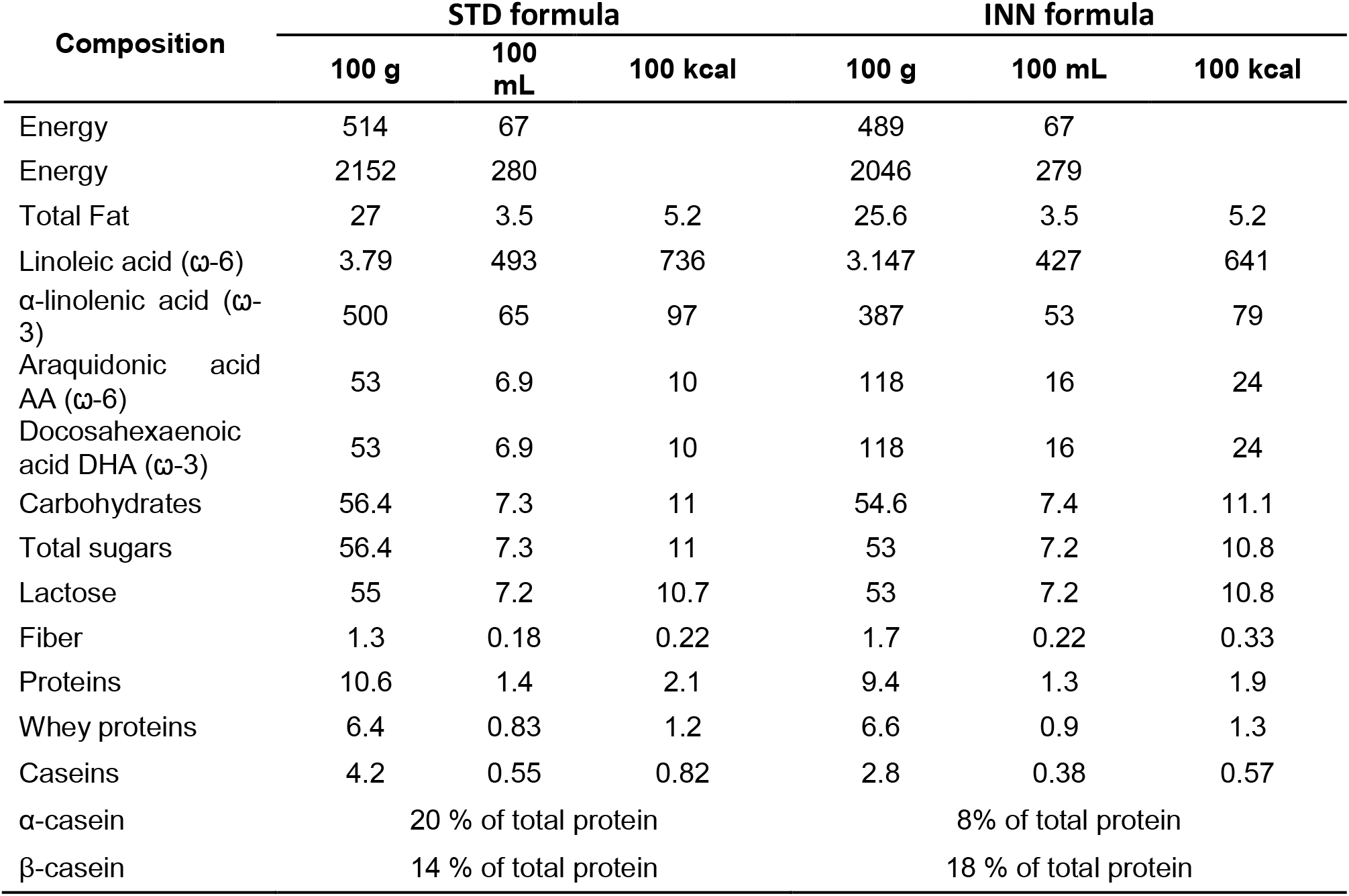
Nutritional composition of the standard infant formula (STD) and study formula (INN).

| Composition | STD formula |  |  | INN formula |  |  |
| --- | --- | --- | --- | --- | --- | --- |
|  | 100 g | 100 mL | 100 kcal | 100 g | 100 mL | 100 kcal |
| Energy | 514 | 67 |  | 489 | 67 |  |
| Energy | 2152 | 280 |  | 2046 | 279 |  |
| Total Fat | 27 | 3.5 | 5.2 | 25.6 | 3.5 | 5.2 |
| Linoleic acid (ω-6) | 3.79 | 493 | 736 | 3.147 | 427 | 641 |
| α-linolenic acid (ω-3) | 500 | 65 | 97 | 387 | 53 | 79 |
| Araquidonic acid AA (ω-6) | 53 | 6.9 | 10 | 118 | 16 | 24 |
| Docosahexaenoic acid DHA (ω-3) | 53 | 6.9 | 10 | 118 | 16 | 24 |
| Carbohydrates | 56.4 | 7.3 | 11 | 54.6 | 7.4 | 11.1 |
| Total sugars | 56.4 | 7.3 | 11 | 53 | 7.2 | 10.8 |
| Lactose | 55 | 7.2 | 10.7 | 53 | 7.2 | 10.8 |
| Fiber | 1.3 | 0.18 | 0.22 | 1.7 | 0.22 | 0.33 |
| Proteins | 10.6 | 1.4 | 2.1 | 9.4 | 1.3 | 1.9 |
| Whey proteins | 6.4 | 0.83 | 1.2 | 6.6 | 0.9 | 1.3 |
| Caseins | 4.2 | 0.55 | 0.82 | 2.8 | 0.38 | 0.57 |
| α-casein | 20 % of total protein | | | 8% of total protein | | |
| β-casein | 14 % of total protein | | | 18 % of total protein | | |

